# Association of depression screening with diagnostic and treatment-related outcomes among youth

**DOI:** 10.1101/2021.04.29.21256334

**Authors:** Kira E. Riehm, Emily Brignone, Elizabeth A. Stuart, Joseph J. Gallo, Ramin Mojtabai

## Abstract

**Background and Objectives:** The goals of depression screening, which is universally recommended in primary care settings in the U.S., are to identify adolescents with depression and connect them to treatment. However, little is known about how depression screening affects the likelihood of being diagnosed with a mental disorder or accessing mental health care over time.

**Methods:** This longitudinal cohort study used insurance claims data from 57,732 adolescents who had at least one routine well-visit between 2014 and 2017. Using propensity score matching, we compared adolescents who were screened for depression to similar adolescents who were not screened for depression during the well-visit. Diagnostic and treatment-related outcomes were examined over 6-month follow-up and included depression diagnoses, mood-related diagnoses, antidepressant prescriptions, any mental health-related prescriptions, and psychotherapy. We also examined heterogeneity of associations by sex.

**Results:** Compared to adolescents who were not screened for depression, adolescents screened for depression were 30% more likely to be diagnosed with depression (RR=1.30, 95% CI=1.11-1.52) and 17% more likely to receive a mood-related diagnosis (RR=1.17, 95% CI=1.08-1.27), but were not more likely to be treated with an antidepressant prescription (RR=1.11, 95% CI=0.82-1.51), any mental health prescription (RR=1.15, 95% CI=0.87-1.53), or psychotherapy (RR=1.13, 95% CI=0.98-1.31). In general, associations were stronger among females.

**Conclusions:** Adolescents who were screened for depression during a well-visit were more likely to receive a diagnosis of depression or a mood-related disorder in the six months following screening. Future research should explore methods for increasing access to treatment and treatment uptake following screening.

**Clinical Trial Registration (if any):** N/A

**Table of Contents Summary:** Insurance claims data were used to explore associations between depression screening during routine well-visits and depression diagnoses, psychiatric prescriptions, and psychotherapy among adolescents.

**What’s Known on This Subject:** Depression screening is increasingly viewed as a key strategy for addressing depression among adolescents. To inform clinical guidelines, the United States Preventive Services Task Force has called for research to examine the diagnostic and treatment-related outcomes of depression screening.

**What This Study Adds:** In a sample of 57,732 adolescents, adolescents who were screened for depression during a well-visit were more likely to receive a depression or mood-related diagnosis over 6-month follow-up, but were not more likely to be treated with medication or psychotherapy.

## INTRODUCTION

Depression is a leading cause of morbidity and functional impairment among adolescents.^1^ Left untreated, adolescent depression can have adverse consequences for well-being in adulthood, including chronicity of depressive symptoms, onset of other mental health disorders, incidence of somatic conditions, and premature mortality.^2-5^ Accumulating evidence suggests that the prevalence of major depressive episodes and depressive symptoms has increased among adolescents over the past decade in the U.S.^6,7^ Together, the negative outcomes associated with depression and rising rates over time demand action to reduce the burden of this mental health problem among adolescents.

The accurate identification of depressive symptoms is a prerequisite to treatment. However, prior studies have found that primary care providers may fail to identify up to two-thirds of adolescents with depression.^8,9^ Screening has been proposed as one method of increasing the detection of depression, which could in turn connect a greater number of patients to mental health care and lead to improved health outcomes.^10^ Universal depression screening is currently recommended in primary care settings for adolescents by the U.S. Preventive Services Task Force (USPSTF),^11^ but this recommendation is controversial, with some researchers pointing to a lack of evidence for improved outcomes following screening.^10,12^

At present, there are no randomized controlled trials (RCTs) that compare depression screening to usual care in primary care settings among adolescents.^12^ Although RCTs are considered the gold standard of evidence for assessing the effects of interventions on health,^13^ an RCT of depression screening in primary care, comparing screened to unscreened adolescents, may not be considered ethically sound because of the universal USPSTF recommendation.^11^ Instead, evidence for depression screening is often inferred from studies of collaborative care or education programs for providers,^14-17^ which are unlikely to generalize to primary care settings where such care or education are unavailable. To better inform clinical guidelines, decision makers have called for research to examine the longitudinal outcomes of depression screening.^11^

In the absence of evidence from RCTs, the application of causal inference methods to observational data provides an important opportunity to estimate the effect of depression screening on outcomes among adolescents. Because physicians may be more likely to screen adolescents they suspect are depressed,^18^ to compare the outcomes of screened and non-screened adolescents, we used propensity scores to account for characteristics associated with being screened in the first place. The objectives of this study were (1) to examine the prospective association of being screened for depression during routine well-visits with diagnostic and treatment-related outcomes among adolescents and (2) to determine if associations varied by sex. We hypothesized that adolescents screened for depression would be more likely to receive a diagnosis of depression and be treated for it in the six months following screening, compared to adolescents not screened for depression.

## METHODS

### Participants

Healthcare claims and pharmacy claims data were obtained from Highmark Health, a non-profit healthcare organization.^19^ The data include Blue Cross Blue Shield (BCBS) members with commercial (large group, small group, and participants who acquired individual or small group insurance through Affordable Care Act Marketplace) or Medicare Advantage insurance. The largest membership is in Pennsylvania, West Virginia, and Delaware, where Highmark Health is the BCBS insurer; however, members live across the U.S. due to large national accounts.

This study used a retrospective cohort design. Participants who met the following criteria were included: (1) had a well-visit (V20.2, V70.0, Z00.121, Z00.129, Z00.00, Z00.01, 99384, 99394, 99385, or 99395) in primary care or general pediatric settings after January 1st, 2014 and before December 31st, 2017 (hereafter referred to as the index well-visit); (2) were continuously enrolled for at least six months before and two years after the index well-visit; and (3) were aged 12-18 (the age group that the USPSTF guidelines apply to) at the time of the index well-visit. For participants with more than one well-visit during the study period, we selected the first well-visit to maximize available follow-up time. In line with other studies,^20^ we excluded adolescents who had a diagnosis of depression, an anti-depressant prescription, or psychotherapy treatment in the six months prior to the index well-visit. Because our interest was in short-term outcomes following depression screening, we selected a follow-up period that extended for six months after the index well-visit.

Of the 281,463 adolescents with a well-visit recorded between 2014 and 2017, we included 57,732 in our propensity score-matched sample; a flow diagram for participant selection is displayed in Supplementary Figure 1.

### Measures

#### Depression Screening

Depression screening was defined according to a combination of ICD-9, ICD-10, Current Procedural Terminology (CPT), and Healthcare Common Procedure Coding System (HCPCS) codes. Similar to a prior study,^20^ adolescents were coded as having been screened for depression during the index well-visit if at least one of the following codes was recorded: G0444, G8510, 96127, 99420, V79.0, and/or Z13.89. A description of each code, and the proportion of well-visits with each code recorded, is presented in Supplementary Table 1.

#### Depression Diagnosis

Because general medical providers tend to use non-specific mood diagnostic categories to identify depressive disorders,^21^ two diagnostic outcomes were used in this study: a narrower diagnosis of depression, and a broader definition of any mood-related diagnosis. We defined these according to peer-reviewed, validated definitions where available; if not, we used definitions from prior studies. Depression diagnoses were identified by ICD-9 codes 296.20-296.25, 296.30-296.35, 300.4, 309.0, 309.1, 309.28 and 311 and ICD-10 codes F31.3-F31.6, F32.0-32.9, F33.0-33.3, F33.8, F33.9, F34.1, F34.8, F34.9, F38.0, F38.1, F38.8, F39, F41.2, and F99.^22^ Mood-related diagnoses were identified with ICD-9 codes 296, 300, 307-309, 311, and 313 and ICD-10 codes F30-F48, F93, and F99.^17^

#### Depression Treatment

The treatment-related outcomes were ascertained from CPT codes and included medication treatment as well as psychotherapy. We separately examined antidepressant medications and any mental health medication (antidepressants, mood-stabilizers, anti-anxiety medications, anti-psychotic medications, and medications to treat attention-deficit hyperactivity disorder); a list of medications included is provided in Supplementary Table 2. A list of the CPT codes included to identify psychotherapy is in Supplementary Table 3.

#### Covariates

We selected characteristics likely to be associated with both selection for depression screening and depression outcomes based on existing literature.^18,20^ Age (years) and sex (male and female) were obtained from claims data. Race/ethnicity (Asian, Black, Hispanic/Latino, other, and White) was estimated using the *wru* R package, which applies Bayesian methods to generate predicted probabilities for each race/ethnicity category for a given person based on geolocation and other individual characteristics.^23^ Rurality was defined using Rural-Urban Commuting Area codes from 2010 (the most recent available), which are provided at the ZIP level; these codes were aggregated into three categories (urban, large rural city/town, and small/isolated rural town). Prior emergency health services use was defined as the count of encounters with emergency or urgent care services in the six months prior to the index well-visit. Prior routine health services use was defined as the count of encounters with outpatient, primary care, and preventive services in the six months prior to the index well-visit. Physical health comorbidities were defined by the count of encounters in the six months prior to the index well-visit where an ICD code for a given category of the Charlson Comorbidity Index was recorded. Data for ZIP-level median household income were obtained from the American Community Survey from 2006-2010.^24^ The medical specialties of the health care providers performing the well-visits were aggregated into three categories (child- or adolescent-specific, family-specific, and other primary care).

### Statistical Analysis

#### Propensity Score Estimation

Our objective was to estimate the effect of being screened for depression, compared to not being screened for depression, on diagnostic and treatment-related outcomes. To this end, we used propensity score matching to account for confounding of the association between screening and outcomes by the covariates listed above. Propensity scores are a causal inference technique that can be applied to observational data to emulate some qualities of RCTs and estimate treatment effects when full-scale RCTs are not feasible.^25^ We used nearest neighbor matching without replacement, selecting three unscreened adolescents for each screened adolescent. Matches were selected based on a propensity score estimated using a logistic regression model with depression screening as the dependent variable and the covariates as independent variables. To account for hypothesized moderation by sex, we included interaction terms between sex and each covariate in the propensity score model and used exact matching for sex.^26^ Figure 1 displays standardized mean differences (SMDs) for all covariates before and after matching. After matching, SMDs for all covariates were less than 0.1, indicating that the matching procedure improved balance between screened and unscreened adolescents on observed covariates. The matched sample (n = 57,732) was composed of 14,433 screened and 43,299 unscreened adolescents.

**Figure 1.**
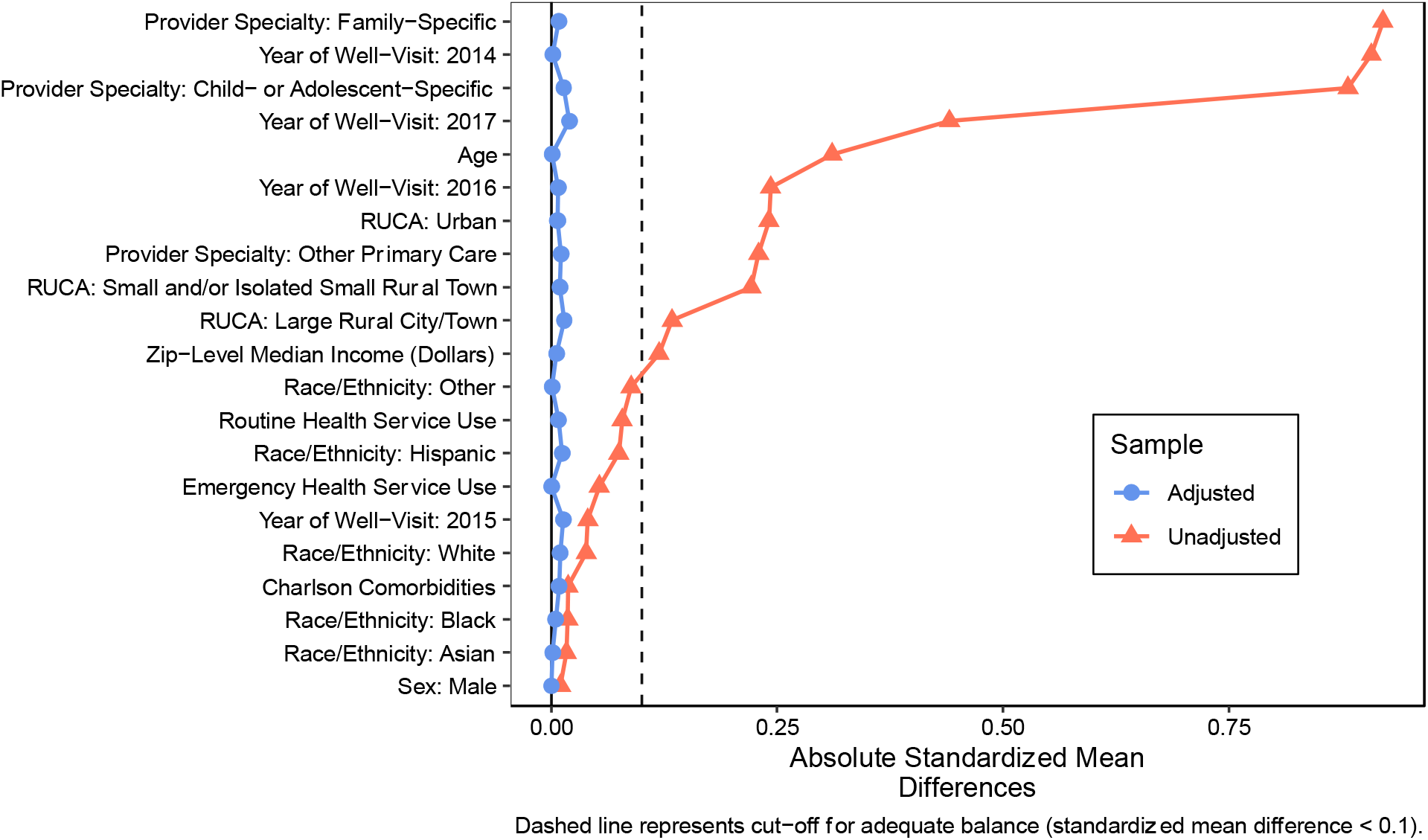
Plot of absolute standardized mean differences for covariates, before (pink) and after (blue) propensity score matching.

#### Descriptive Characteristics

We examined descriptive statistics in both the complete sample and the propensity score-matched sample. We also examined changes in the proportion of adolescents screened over time.

#### Association of Depression Screening with Diagnostic and Treatment-Related Outcomes

To estimate the effect of depression screening on each outcome, we estimated a series of log-binomial regression models using the matched sample with depression screening as the independent variable. To determine if the effect of depression screening on each outcome varied by sex, we estimated the same series of models including an interaction term between depression screening and sex. Coefficients for the depression screening variable were exponentiated for interpretation as risk ratios (RRs) representing the average effect of screening among those who were screened (i.e., the average treatment effect among the treated). Standard errors in all models were estimated using cluster-robust variance estimates to account for matched clusters.

#### Sensitivity Analysis

Our study period coincided with the implementation of ICD-10 in the U.S. on October 1, 2015. To test the sensitivity of our findings to the change in coding system, we re-estimated our models with an interaction term between depression screening and an indicator variable for whether an index well-visit occurred before or after October 1, 2015. We also tested the sensitivity of significant parameter estimates to unmeasured confounding by calculating e-values, which convey the minimum strength of association that an unmeasured confounder would need to have with both the depression screening and each outcome to fully explain away associations.^27^

Statistical significance was assessed at p<.05. All analyses were conducted using R (R studio version 1.2.5042; R version 4.0.0). This study was reviewed by the Johns Hopkins Institutional Review Board and was determined to be human subjects research that meets criteria for exemption under category four (i.e., secondary analysis of existing, de-identified data).

## RESULTS

### Descriptive Characteristics

Descriptive characteristics for the complete and matched samples are displayed in Table

1. The sample of 248,354 adolescents had a mean age of 14.26 years (standard deviation [SD]=2.06) and 121,432 (48.9%) were female. In the 6-month period following the index well-visit, 4,730 (1.9%) adolescents received a depression diagnosis, 13,591 (5.5%) received a mood-related diagnosis, 1,330 (0.5%) were treated with antidepressants, 5,182 (2.1%) were treated with a prescription for any mental health condition, and 6,394 (2.6%) were treated with psychotherapy.

The percentage of adolescents screened for depression during the index well-visit increased across the study period (Figure 2). In 2014, approximately 2.0% of adolescents were screened for depression during the index well-visit; in contrast, of adolescents who had their index well-visit in 2017, 13.6% were screened.

**Figure 2.**
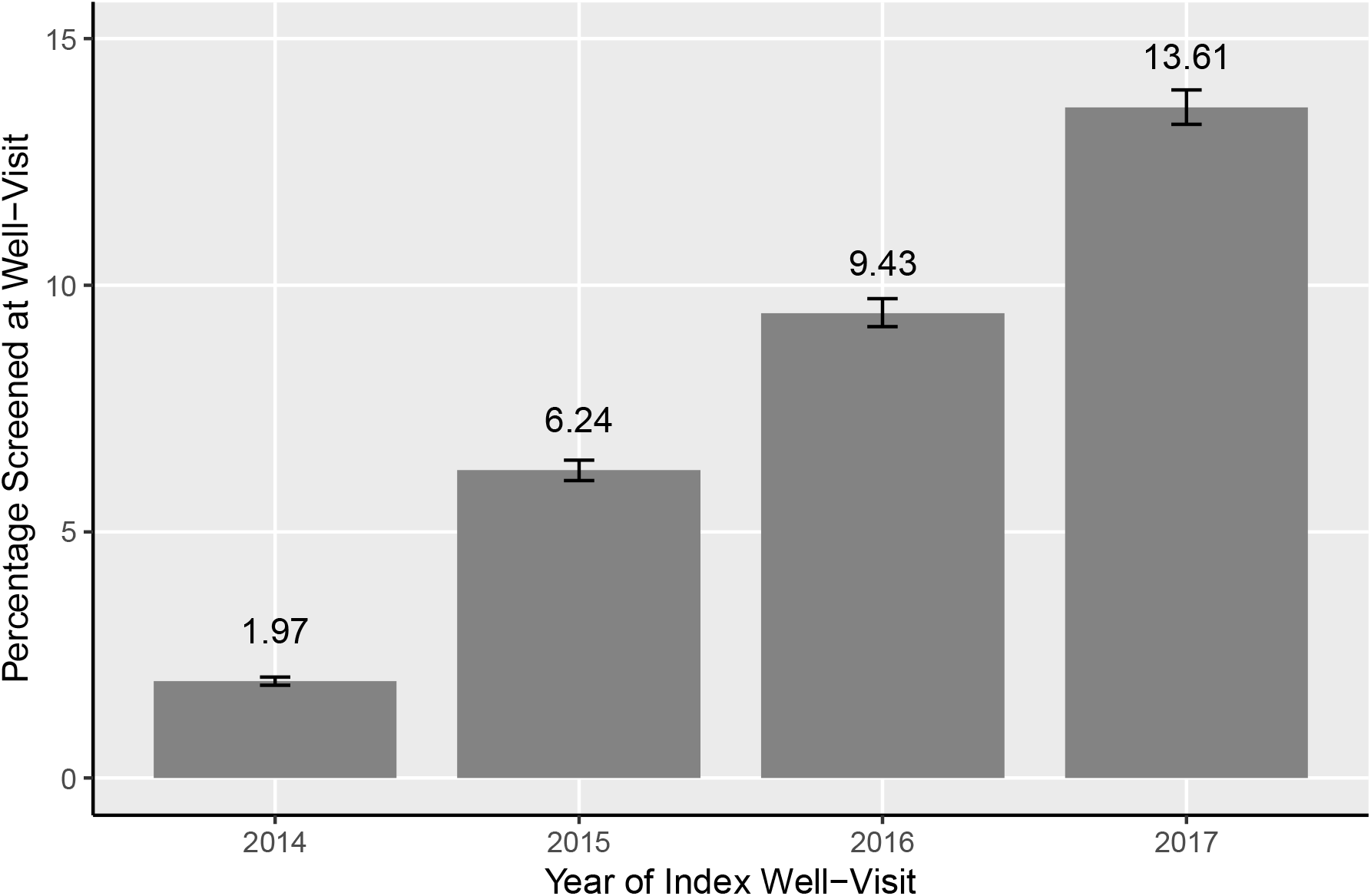
Percentage of adolescents screened for depression at the index well-visit from 2014-2017.

### Association of Depression Screening with Diagnostic and Treatment-Related Outcomes

Results of the propensity score matched analyses are displayed in Table 2. Compared to adolescents not screened for depression, those who were screened during the index well-visit were 30% more likely to be diagnosed with depression (RR=1.30, 95% CI=1.11-1.52) and 17% more likely to receive a mood-related diagnosis (RR=1.17, 95% CI=1.08-1.27) across six-month follow-up. Being screened for depression was not associated with antidepressant prescriptions (RR=1.11, 95% CI=0.82-1.51), any mental health prescriptions (RR=1.15, 95% CI=0.87-1.53), or psychotherapy (RR=1.13, 95% CI=0.98-1.31).

In stratified analyses (Table 2), females who were screened for depression were significantly more likely to be diagnosed with depression (RR=1.42, 95% CI=1.17-1.71), to receive a mood-related diagnosis (RR=1.26, 95% CI=1.12-1.40), and to be treated with psychotherapy (RR=1.22, 95% CI=1.05-1.43), compared to females not screened for depression. These same associations were not significant among males (depression diagnosis: RR=1.12, 95% CI=0.87-1.44; mood-related diagnosis: RR=1.06, 95% CI=0.93-1.22; psychotherapy: RR=1.02, 95% CI=0.84-1.23). However, interactions terms between depression screening and sex were not statistically significant for any outcome.

### Sensitivity Analysis

Interaction terms between depression screening and an indicator variable for whether an index well-visit occurred before or after October 1, 2015 were not significant in models for any outcome, indicating that our results were unlikely to have been influenced by the implementation of ICD-10. E-values for significant parameter estimates are presented in Supplementary Table 4. The observed risk ratios of 1.30 and 1.17 for depression diagnoses and mood-related diagnoses could be explained away by an unmeasured confounder that was associated with both depression screening and these outcomes by a risk ratios of 1.92 and 1.62, respectively, above and beyond the measured confounders, but weaker confounding could not do so. Given other aspects of our study design, namely the exclusion of those with recent depression diagnoses and/or depression treatment, we interpret these e-values as indicating that unmeasured confounding is possible but unlikely.

## DISCUSSION

In this study, we found that adolescents who were screened for depression during a well-visit were more likely to subsequently be diagnosed with depression or a mood-related condition over six-month follow-up. However, they were not more likely to fill an antidepressant or other mental health prescription, nor receive psychotherapy. Associations between depression screening and outcomes were generally stronger among females compared to males.

Our study builds on prior findings^17,28-31^ by using rigorous causal inference methods to examine longitudinal outcomes of depression screening in a large, population-based sample of adolescents. Our results suggest that depression screening may increase identification of depression and other behavioral health conditions but may not reduce unmet need for care. The USPSTF recommendation for depression screening states that “adequate systems should be in place to ensure accurate diagnosis, effective treatment, and appropriate follow-up”^11^ following screening. However, others have noted that primary care settings rarely have these systems in place due to scarce treatment resources and inadequate funding to support training of providers.^32^ The scope of our study precludes examining factors that may have played a role in lack of treatment uptake, but broadly, our findings suggest that structural barriers may limit successful engagement in treatment following screening. We also observed that the proportion of adolescents screened for depression during the index well-visit increased over time. Altogether, this suggests that over time, increasing number of adolescents are identified as depressed but go untreated, which highlights the urgency of addressing barriers to care.

We observed some evidence of heterogeneity by sex; associations between depression screening and outcomes tended to be stronger among female adolescents compared to males. A well-replicated finding is that males tend to report greater mental health stigma, which serves as a substantial deterrent to help-seeking.^33^ In the context of depression screening, males may be more reluctant than females to engage in follow-up care. Another explanation is that males may express depression differently from females, with greater symptoms of anger, irritability, and emotional numbness.^34^ These symptoms are not commonly assessed with generic screening tools and may not be recognized as depression.

A number of strategies have been introduced to increase mental health services utilization among adolescents. Prior studies have noted the central role that parents play in the depression screening process; in particular, many primary care providers perceive parental resistance to diagnosis and negative attitudes toward treatment to be key barriers to adolescent entry to treatment.^35^ Instead of viewing parental attitudes as a barrier to care, active approaches that provide psychoeducation to parents and involve them in routine treatment decisions are likely to be more effective at ensuring treatment needs are met.^36^ Preliminary studies of “warm hand-offs,” where an adolescent is introduced to a behavioral health provider at the time of referral, have been found to increase the likelihood of attendance at follow-up appointments;^37,38^ however, these models may only be feasible in integrated settings where behavioral health providers are on-site. Other strategies have focused on integration of psychosocial screening tools into electronic health records and the use of computer-based decision support systems, which can improve workflow and streamline referral processes.^30,39-41^ Broader policy-based efforts are also critical for increasing the supply of mental health providers, as well as employing consultation models whereby psychiatrists can guide the management of adolescent mental health problems by primary care providers.^42^ Finally, the COVID-19 pandemic has resulted in rapid roll-out of telehealth services, and there is hope that these services can help overcome geographic and financial barriers to care.^43^

Some limitations of this study should be noted. The sample used in this study is based primarily in the Eastern U.S. and may not generalize to the country as a whole. We used health insurance claims data, which are limited in the amount of information they can provide about a given medical encounter. For example, we were unable to access the results of depression screening procedures. Although we used validated exposure and outcome definitions where possible, the accuracy of claims data in reflecting certain procedures and health outcomes remains an area of active research.^44^ Adolescents in our sample may have accessed mental health treatment not covered by insurance, which would not have been captured by claims data. Finally, propensity scores cannot guarantee balance on unobserved confounders, though our sensitivity analyses suggested that residual confounding is unlikely. On the other hand, strengths of this study include the use of a large, longitudinal sample to examine rare outcomes and the application of rigorous causal inference methods to estimate effects.

In summary, this study found that adolescents who were screened for depression during a well-visit were more likely to receive a diagnosis of depression or a mood-related disorder in the six months following screening; however, they were not more likely to be treated with prescription medication or psychotherapy. Associations between screening and outcomes were generally stronger among females compared to males. Future studies should examine long-term health outcomes of depression screening, as well as the implementation of strategies to increase treatment uptake among adolescents.

## Data Availability

These data are covered under a Data Use Agreement between the Johns Hopkins Bloomberg School of Public Health and Highmark Health.

## Abbreviations

(BCBS): Blue Cross Blue Shield
(CPT): Current Procedural Terminology
(HCPCS): Healthcare Common Procedure Coding System
(ICD): International Classification of Diseases
(RCT): Randomized controlled trial
(US): Relative risk (RR); United States
(USPSTF): United States Preventive Services Task Force

---

**Table 1.** Descriptive characteristics for complete and matched samples, N (%).

| Variable | Complete Sample<br>(n=248,354) | Matched Sample (n=57,732) |  |
| --- | --- | --- | --- |
|  |  | Not Screened<br>(n=43,299) | Screened<br>(n=14,433) |
| Covariates |  |  |  |
| Age<sup>a</sup> | 14.26 (2.06) | 13.68 (1.98) | 13.69 (1.96) |
| Sex |  |  |  |
| Female | 121,432 (48.9%) | 21,375 (49.4%) | 7,125 (49.4%) |
| Male | 126,922 (51.1%) | 21,924 (50.6%) | 7,308 (50.6%) |
| Race/Ethnicity<sup>a,b</sup> |  |  |  |
| Asian | 0.02 (0.11) | 0.03 (0.12) | 0.03 (0.12) |
| Black | 0.07 (0.17) | 0.07 (0.18) | 0.08 (0.18) |
| Hispanic/Latino | 0.07 (0.18) | 0.05 (0.16) | 0.05 (0.17) |
| Other | 0.02 (0.04) | 0.01 (0.03) | 0.01 (0.04) |
| White | 0.82 (0.28) | 0.83 (0.26) | 0.83 (0.27) |
| Rurality |  |  |  |
| Urban | 199,569 (80.4%) | 37,914 (87.6%) | 12,670 (87.8%) |
| Large Rural City/Town | 30,039 (12.1%) | 3,883 (9.0%) | 1,237 (8.6%) |
| Small/Isolated Rural Town | 18,746 (7.5%) | 1,502 (3.5%) | 526 (3.6%) |
| Year of Index Well-Visit |  |  |  |
| 2014 | 117,743 (47.4%) | 6,968 (16.1%) | 2,314 (16.0%) |
| 2015 | 54,054 (21.8%) | 10,362 (23.9%) | 3,373 (23.4%) |
| 2016 | 40,024 (16.1%) | 11,467 (26.5%) | 3,775 (26.2%) |
| 2017 | 36,533 (14.7%) | 14,502 (33.5%) | 4,971 (34.4%) |
| Emergency Health Services Use<sup>a</sup> | 0.18 (0.52) | 0.21 (0.58) | 0.21 (0.54) |
| Routine Health Services Use<sup>a</sup> | 0.85 (1.84) | 0.72 (1.50) | 0.73 (1.61) |
| Physical Health Comorbidities<sup>a</sup> | 0.12 (1.50) | 0.09 (0.65) | 0.10 (0.84) |
| ZIP-Level Median Income<sup>a</sup> | 55,886.52 (19,086.55) | 58,219.29 (20,148.41) | 58,107.51 (19,806.49) |
| Provider Specialty |  |  |  |
| Child- or Adolescent-Specific | 159,647 (64.3%) | 38,984 (90.0%) | 12,935 (89.6%) |
| Family-Specific | 67,957 (27.4%) | 2,640 (6.1%) | 909 (6.3%) |
| Other Primary Care | 20,750 (8.4%) | 1,675 (3.9%) | 589 (4.1%) |
| Outcomes |  |  |  |
| Depression Diagnosis<sup>c</sup> | 4,730 (1.9%) | 761 (1.8%) | 329 (2.3%) |
| Mood-Related Diagnosis<sup>c</sup> | 13,591 (5.5%) | 2,426 (5.6%) | 948 (6.6%) |
| Antidepressant Prescription<sup>c</sup> | 1,330 (0.5%) | 224 (0.5%) | 83 (0.6%) |
| Any Mental Health Prescription<sup>c</sup> | 5,182 (2.1%) | 972 (2.2%) | 374 (2.6%) |
| Psychotherapy<sup>c</sup> | 6,394 (2.6%) | 1,295 (3.0%) | 488 (3.4%) |

Notes: <sup>a</sup>Reported as mean (standard deviation). <sup>b</sup>Race/ethnicity is a predicted probability based on the *wru* package in R. <sup>c</sup>Cells report the count and proportion of adolescents with the outcome. Bold font indicates statistical significance.

**Table 2.** Propensity-score adjusted association of depression screening with diagnostic and treatment-related outcomes, in the matched sample (n=57,732) and stratified by sex.

| Outcome | Main Association of Depression Screening |  | With Interaction Term Between Depression Screening and Sex |  |  |  | p-value for difference |
| --- | --- | --- | --- | --- | --- | --- | --- |
|  | RR | 95% CI | Females |  | Males |  |  |
|  |  |  | RR | 95% CI | RR | 95% CI |  |
| Depression Diagnosis | 1.30 | 1.11, 1.52 | 1.42 | 1.17, 1.71 | 1.12 | 0.87, 1.44 | 0.172 |
| Mood-Related Diagnosis | 1.17 | 1.08, 1.27 | 1.26 | 1.12, 1.40 | 1.06 | 0.93, 1.22 | 0.070 |
| Antidepressant Prescription | 1.11 | 0.82, 1.51 | 1.28 | 0.89, 1.84 | 0.85 | 0.51, 1.44 | 0.286 |
| Any Mental Health Prescription | 1.15 | 0.87, 1.53 | 1.23 | 0.99, 1.53 | 1.10 | 0.92, 1.32 | 0.633 |
| Psychotherapy | 1.13 | 0.98, 1.31 | 1.22 | 1.05, 1.43 | 1.02 | 0.84, 1.23 | 0.175 |

Notes: Bold font indicates statistical significance.

