## Supplementary files for "Association of depression screening with diagnostic and treatment-related outcomes among youth"

**Supplementary Figure 1.** Participant flow diagram.

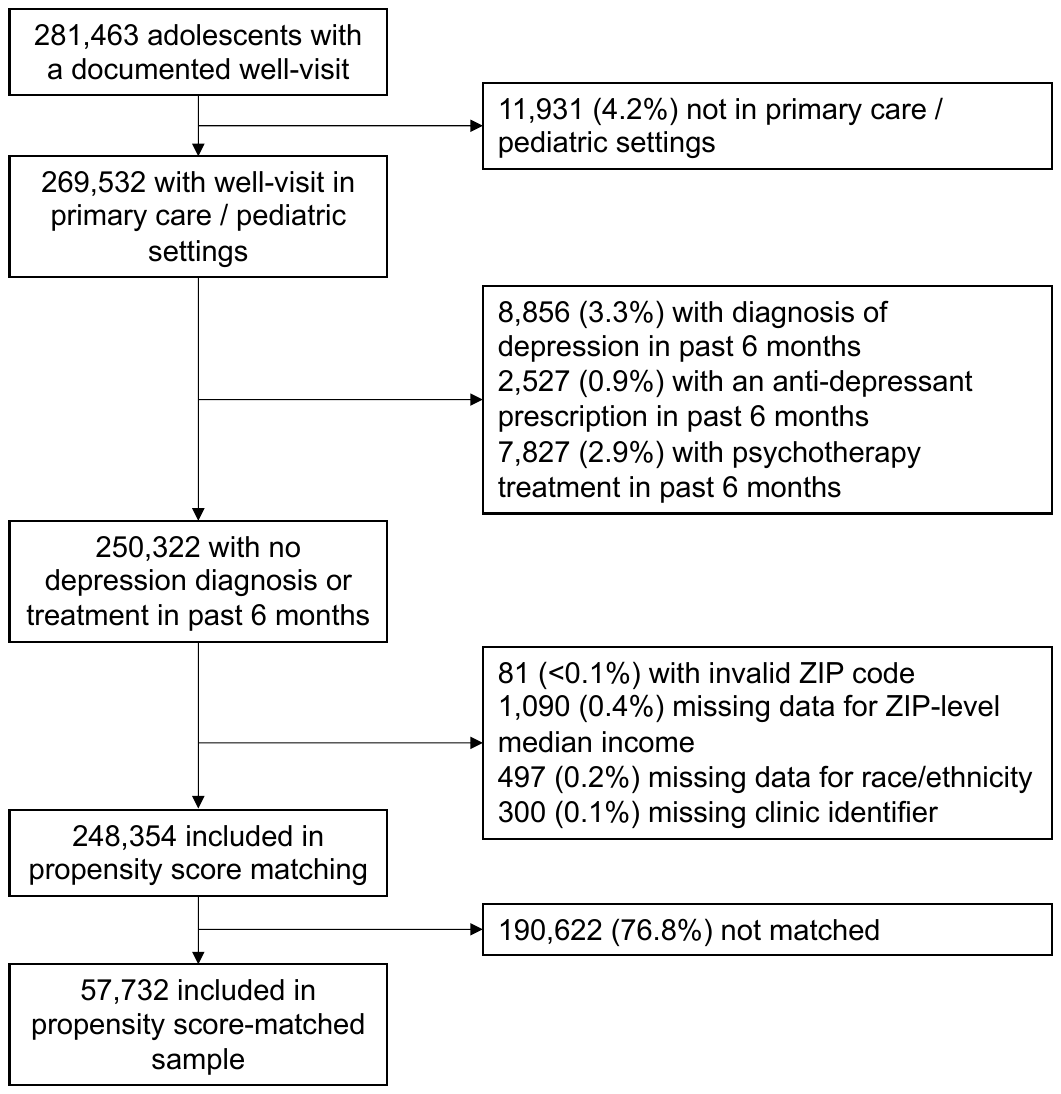

**Supplementary Table 1.** Description of depression screening codes and n (%) of well-visits where each code was reported.

| **Code** | **Description** | **N (%)** |
| --- | --- | --- |
| G0444 | Annual depression screening | 3,067 (21.3) |
| G8510 | Screening for depression is documented as negative, a follow-up plan is not required | 11 (0.1) |
| 96127 | Brief emotional/behavioral assessment (eg, depression inventory, attention-deficit/hyperactivity disorder [ADHD] scale), with scoring and documentation, per standardized instrument | 2,504 (17.3) |
| 99420 | Administration and interpretation of health risk assessment instrument | 461 (3.2) |
| V79.0 | Screening for depression | 709 (4.9) |
| Z13.89^a^ | Encounter for screening for other disorder | 1,359 (9.4) |
| Two or more of the above codes | N/A | 6,322 (43.8) |
| Notes: ^a^This code is identified by the American Academy of Family Physicians as coding for depression screening (https://www.aafp.org/fpm/2016/0900/p40.html). | | |

**Supplementary Table 2.** List of medications included in outcome definitions for anti-depressant medications and any mental health prescription.

| **Category** | **Specific Medication Names** |
| --- | --- |
| Selective serotonin reuptake inhibitors (SSRIs) | Citalopram (Celexa)  Escitalopram (Lexapro)  Fluoxetine (Prozac, Sarafem)  Fluvoxamine (Luvox)  Olanzapine/fluoxetine  Paroxetine (Paxil, Pexava)  Sertraline (Zoloft)  Fluoxetine/olanzapine (Symbyax) |
| Serotonin and norepinephrine reuptake inhibitors (SNRIs) | Desvenlafaxine (Khedezla, Pristiq)  Duloxetine (Cymbalta)  Levomilnacipran (Fetzima)  Venlafaxine (Effexor, Effexor XR, Venlafaxine) |
| Monoamine oxidase inhibitors (MAOIs) | Iscorboxazid (Marplan)  Phenelzine (Nardil)  Selegiline transdermal (Emsam)  Tranylcypromine (Parnate) |
| Norepinephrine and dopamine reuptake inhibitors (NDRIs) | Bupropion (Wellbutrin, Aplenzin, Forfivo XL) |
| Atypical antidepressants | Mirtazapine (Remeron)  Trazodone  Vilazodone (Viibryd)  Vortioxetine (Trintellix) |
| Tri- and tetracyclic antidepressants | Amitriptyline (Elavil)  Amoxapine  Chlordiazepoxide/amitriptyline (Limbitrol, Limbitrol DS)  Clomipramine  Desipramine (Norpramin)  Doxepine  Imipramine (Tofranil, Tofranil PM)  Maprotiline  Nortiptyline (Pamelor)  perphenazine/amitriptyline  Protriptyline (Vivactil)  Trimipramine (Surmontil) |
| Mood stabilizers | Carbamazepine (Tegretol, Epitol)  Divalproex, valproic acid or valproate (Depakene, Epival)  Lamotrigine (Lamictal)  Lithium- lithium carbonate or lithium citrate (Carbolith, Duralith, Lithane)  Oxcarbazepine (Trileptal) |
| Benzodiazepines | Alprazolam (Xanax, Xanax XR, Niravam)  Clobazam (Onfi)  Clonazepam (Klonopin)  Clorazepate (Tranxene)  Chlordiazepoxide (Librium)  Diazepam (Valium, Diastat Acudial, Diastat)  Estazolam  Lorazepam (Ativan)  Oxazepam  Temazepam (Restoril)  Triazolam (Halcion) |
| Antipsychotics | Aripiprazole (Abilify, Aristada)  Asenapine (Saphris)  Chlorpromazine  Fluphenazine  Haloperidol  Lurasidone (Latuda)  Olanzapine (Zyprexa)  Paliperidone (Invega)  Perphenazine  Quetiapine (Seroquel)  Risperidone (Risperdal)  Ziprasidone (Geodon) |
| ADHD Medications^a^ | Adderall  Amphetamine  Desoxyn  Phenopromin  Amfetamine  Phenamine  Centramina  Fenamine  Levoamphetamine  Dexamfetamine  Dexamphetamine  Dexedrine  Dextroamphetamine  DextroStat  Oxydess  Methylamphetamine  Methylenedioxyamphetamine  Methamphetamine  Chloroamphetamine  Metamfetamine  Deoxyephedrine  Desoxyephedrine  Ecstasy  Atomoxetine  Biphentin  Bupropion  Amfebutamone  Zyntabac  Quomen  Wellbutrin  Zyban  Catapres  Clonidine  Klofenil  Clofenil  Chlophazolin  Gemiton  Hemiton  Isoglaucon  Klofelin  Clopheline  Clofelin  Dixarit  Concerta  Daytrana  Methylphenidate  Equasym  Methylin  Tsentedrin  Centedrin  Phenidylate  Ritalin  Duraclon  Elvanse  Focalin  Dexmethylphenidate  Guanfacine  Estulic  Tenex  Kapvay  Lisdexamfetamine  Vyvanse  Medikinet  Metadate  Modafinil  Nexiclon  Quillivant  Strattera |
| Notes: ^a^Obtained from Cortese et al., Cortese S, Adamo N, Del Giovane C, Mohr-Jensen C, Hayes AJ, Carucci S, Atkinson LZ, Tessari L, Banaschewski T, Coghill D, Hollis C. Comparative efficacy and tolerability of medications for attention-deficit hyperactivity disorder in children, adolescents, and adults: a systematic review and network meta-analysis. The Lancet Psychiatry. 2018 Sep 1;5(9):727-38. | |

**Supplementary Table 3.** List of codes used in outcome definition for psychotherapy.

| **Type of Code** | **Codes Included** |
| --- | --- |
| CPT | 90804  90805  90806  90807  90808  90809  90810  90811  90812  90813  90816  90817  90818  90819  90822  90832  90833  90834  90836  90837  90838  90839  90840  90845  90846  90847  90849  90853  90857  90880  96152  96153  96154  96155  99484  99492  99493  99494  0364T  0365T  0366T  0367T  0368T  0369T  0370T  0371T  0372T  0373T  0374T |
| HCPCS | G0409  G0410  G0411  G0502  G0503  G0504  G0507  H0036  H2017  H2018  H2027  H2032  H2033  S9480  S9484  S9485 |

**Supplementary Table 4.** E-values for statistically significant parameter estimates to test sensitivity to unobserved confounding.

| **Outcome** | **Main Association of Depression Screening** | | |
| --- | --- | --- | --- |
|  | **RR** | **95% CI** | **E-Value (Lower Bound of 95% CI)^a^** |
| Depression Diagnosis | **1.30** | **1.11, 1.52** | 1.92 (1.30) |
| Mood-Related Diagnosis | **1.17** | **1.08, 1.27** | 1.62 (1.37) |
| Antidepressant Prescription | 1.11 | 0.82, 1.51 | NA |
| Any Mental Health Prescription | 1.15 | 0.87, 1.53 | NA |
| Psychotherapy | 1.13 | 0.98, 1.31 | NA |
| Notes: Bold font indicates statistical significance. ^a^Calculated according to formulas presented in VanderWeele & Ding, Annals of Internal Medicine, 2017. | | | |
